# Insomnia prehabilitation in newly diagnosed breast cancer patients: protocol for a pilot, multicentre, randomised controlled trial comparing nurse delivered sleep restriction therapy to sleep hygiene education (INVEST trial)

**DOI:** 10.1101/2024.05.30.24308194

**Authors:** Leanne Fleming, Solveiga Zibaite, Simon D. Kyle, Kathleen Boyd, Vivien Green, James Mansell, Beatrix Elsberger, David Young

## Abstract

**Introduction:** Insomnia is a prevalent sleep disorder that negatively impacts daytime functioning and quality of life. Breast cancer patients report higher rates of insomnia and more circadian disruption than other cancer groups. Approximately 50% of patients experience acute insomnia following breast cancer diagnosis, which often persists during cancer treatment and rehabilitation. Sleep Restriction Therapy (SRT) is a clinically effective and tolerable treatment for persistent insomnia in breast cancer survivors. However, SRT has never been tested on patients with early signs of sleep disturbance who are undergoing cancer treatment. The aim of this pilot randomised controlled trial is to explore the feasibility and preliminary effectiveness of nurse delivered SRT for newly diagnosed breast cancer patients with acute insomnia. The trial has been registered on ClinicalTrials.gov (identifier: NCT06294041).

**Methods:** The INVEST (INvestigating the Value of Early Sleep Therapy) trial will recruit 50 newly diagnosed breast cancer patients who meet criteria for acute insomnia. Patients will be recruited from breast cancer results clinics within two Scottish health boards (NHS Grampian and NHS Greater Glasgow and Clyde) and will be block randomised (1:1) to receive nurse delivered SRT or Sleep Hygiene Education (SHE). SRT will be delivered over 4 weekly sessions comprising two face-to-face meetings (either in person or online) and two telephone calls, whereas SHE will be administered in booklet form. Outcomes will be collected at baseline, 6 weeks, and 12 weeks post-randomisation. Primary outcomes in this trial relate to the feasibility of SRT for newly diagnosed breast cancer patients with acute insomnia. Specifically, we will explore (i) rates of patient recruitment and retention, (ii) intervention fidelity, (iii) data collection procedures and outcome measure completion, (iv) intervention acceptability. Secondary outcomes will focus on preliminary evaluation of patient responses to SRT, including insomnia severity, rest-activity rhythms, and mental health.

**Ethics and dissemination:** Ethical approval has been granted by the West of Scotland REC 3 committee (reference 23/WS/0113). Our dissemination plan comprises publishing trial outcomes in high-impact, peer-reviewed journals and on breast cancer charity websites and other patient resources. The outcomes from this pilot trial will also inform the development of a full-scale, multicentre RCT of SRT for acute insomnia in newly diagnosed breast cancer patients. University of Strathclyde is the sponsor (reference: UEC23/52). Protocol version v1.2 4 October 2023.

Strengths and limitations of this study

1. This trial is the first to explore the value of sleep prehabilitation for newly diagnosed breast cancer patients.
2. This will be the first trial to assess the feasibility of delivering SRT during breast cancer treatment, providing valuable insight into its tolerability and preliminary effectiveness.
3. An embedded process evaluation will assess the acceptability of SRT, providing insight into potential optimisation of the intervention and recommendations for enhancing its future scalability and translation within cancer care.
4. Due to the nature of the SRT intervention, nurse therapists and patients cannot be blinded to treatment allocation, increasing the risk of bias.

## INTRODUCTION

Insomnia disorder (ID) is characterised by persistent difficulty with sleep onset, maintenance, and/or early morning awakening, that impacts daytime functioning. It is an important public health problem with substantial medical, psychiatric, and financial ramifications. ID is a causal factor in the evolution and maintenance of physical and mental ill health (1), a risk factor for all-cause mortality (2), and the most commonly reported mental health complaint amongst cancer patients (3, 4). Breast cancer patients report the highest rates of ID and more circadian disruption than other cancer groups (3), which is associated with significantly shorter overall cancer survival, independent of other prognostic factors (5).

Spielman’s diathesis-stress model (1987) of insomnia development explains how the stress of cancer diagnosis precipitates the onset of acute insomnia, then altered rest/activity rhythms, implemented to combat the impact of acute insomnia, perpetuate its transition to ID (6). Our group have previously completed a programme of work exploring this model of insomnia aetiology in breast cancer (7-10). We reported that 46% of newly diagnosed breast cancer patients develop acute insomnia following diagnosis (7). Rates of insomnia remain stable and pervasive for at least 12-months post-diagnosis, indicating that, if not addressed, acute insomnia develops a chronic course (7, 10). We also found that breast cancer patients attempt to compensate for the impact of acute insomnia by engaging in behaviours known to promote the development of ID (i.e., daytime napping, time in-bed extension) (8). Finally, we reported that ID has a significant negative impact on the psychological and physical wellbeing of people with breast cancer (9). Therefore, breast cancer patients would benefit from early intervention for acute sleep disturbance to mitigate its transition to ID.

Sleep Restriction Therapy (SRT) is an established, single-component treatment for ID and a key ingredient within multicomponent Cognitive Behavioural Therapy for insomnia (CBT-I) (11). SRT targets altered rest/activity rhythms, with the aim of increasing homeostatic sleep pressure, over-riding cognitive and physiological arousal, and strengthening circadian control of sleep (11). This is because sleep extension leads to a disparity between sleep opportunity and sleep ability, alters the regularity of the sleep-wake schedule, and potentially introduces irregularity in light exposure. A recent meta-analysis of RCTs demonstrates that SRT significantly improves insomnia severity and sleep continuity in patients with ID (11). Therefore, it offers promise as an early therapeutic approach for acute insomnia.

Our data also show that insomnia interventions (such as SRT) improve concomitant symptoms like anxiety, depression, and fatigue in breast cancer patients (12, 13). In addition, because SRT leads to reduced sleep variability and improved sleep consolidation and quality (14, 15), it may minimise these other prevalent cancer-related side effects as they emerge, so sleep is a promising transdiagnostic therapeutic target for wellbeing more generally. However, whilst SRT may be particularly useful for breast cancer patients with acute insomnia, there are no efficacy studies of single component SRT for acute insomnia in the breast cancer patient population.

### Objectives

Our main objective is to assess the feasibility and preliminary effectiveness of SRT for newly diagnosed breast cancer patients with acute insomnia. Primary outcomes relate to issues of recruitment and retention, intervention fidelity, data collection procedures, completion of outcome measures, and intervention acceptability. Table 1 summarises the schedule of assessments. Table 2 summarises the primary outcomes and corresponding success criteria for moving forward to a full trial, which are based on a similar pilot RCT of breast cancer patients with insomnia (16) and a RCT of SRT in primary care (17). Secondary outcomes relate to the effects of SRT, compared to SHE, on sleep, rest-activity rhythms, and mental health.

**Table 1.**
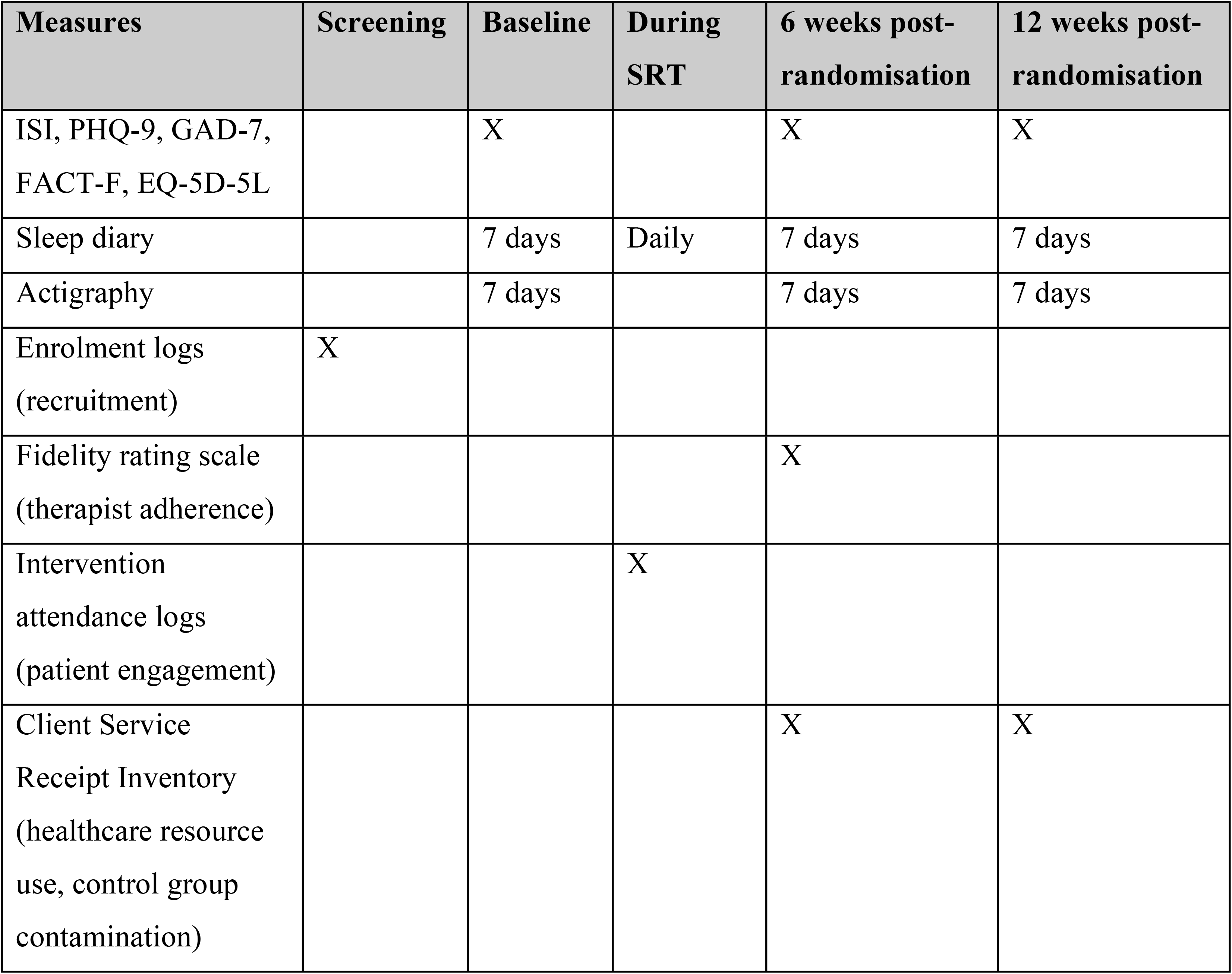
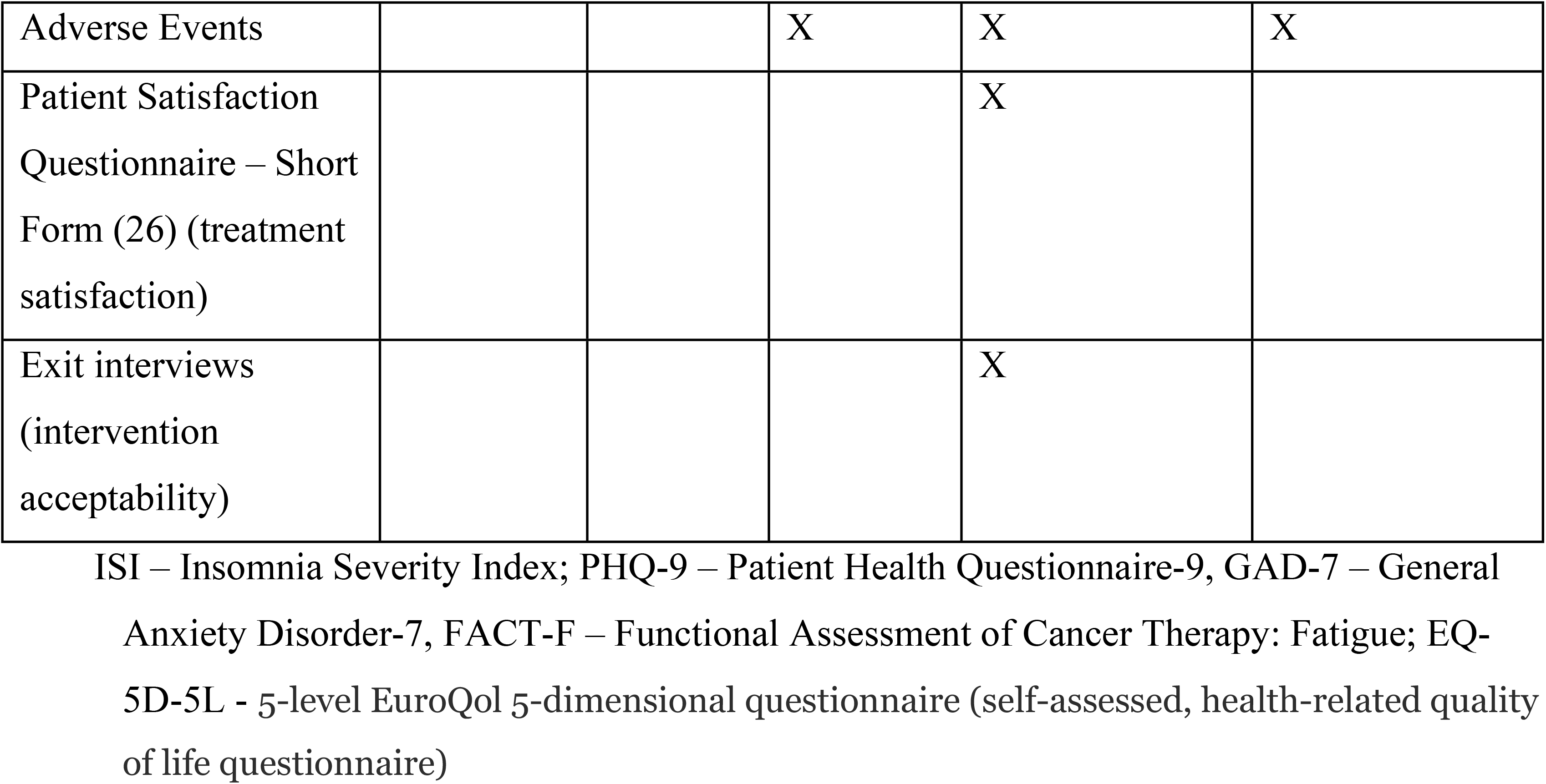
Schedule of Assessments

| Measures | Screening | Baseline | During SRT | 6 weeks post-randomisation | 12 weeks post-randomisation |
| --- | --- | --- | --- | --- | --- |
| ISI, PHQ-9, GAD-7, FACT-F, EQ-5D-5L |  | X |  | X | X |
| Sleep diary |  | 7 days | Daily | 7 days | 7 days |
| Actigraphy |  | 7 days |  | 7 days | 7 days |
| Enrolment logs (recruitment) | X |  |  |  |  |
| Fidelity rating scale (therapist adherence) |  |  |  | X |  |
| Intervention attendance logs (patient engagement) |  |  | X |  |  |
| Client Service Receipt Inventory (healthcare resource use, control group contamination) |  |  |  | X | X |
| Adverse Events |  |  | X | X | X |
| Patient Satisfaction Questionnaire – Short Form (26) (treatment satisfaction) |  |  |  | X |  |
| Exit interviews (intervention acceptability) |  |  |  | X |  |

ISI – Insomnia Severity Index; PHQ-9 – Patient Health Questionnaire-9, GAD-7 – General Anxiety Disorder-7, FACT-F – Functional Assessment of Cancer Therapy: Fatigue; EQ-5D-5L - 5-level EuroQol 5-dimensional questionnaire (self-assessed, health-related quality of life questionnaire)

**Table 2.** Overview of Primary Outcomes and Success Criteria

| **Outcome** | **Quantitative success criteria** | **Quantitative measurement** | **Timepoint(s) of evaluation** | **Qualitative insights (evaluated throughout)** |
| --- | --- | --- | --- | --- |
| **Recruitment and retention rates** | Recruitment - 80% of target achieved | Enrolment logs for all consenting patients. Pre-screen failure logs for patients who meet inclusion criteria but are not enrolled | Screening | Understanding trial implementation and participation / offer of uptake |
|  | Retention - <30% attrition rate | Completion of outcome measures | 12 weeks post randomisation |  |
| **Intervention fidelity** | Nurse therapist adherence - >70% of SRT elements covered | Independent review of recorded SRT sessions and | 6 weeks post randomisation | Understanding nurse therapist and patient perspectives on delivery and |
|  | during sampled sessions | completion of fidelity rating scale |  | receipt of SRT sessions |
|  | Patient engagement - attendance at 2/4 SRT sessions | Intervention attendance logs | During intervention phase |  |
|  | Control group contamination - <10% of SHE patients receive SRT from nurse | Client Service Receipt Inventory | 6 weeks and 12 weeks post randomisation |  |
| **Outcome measure completion** | >70% of questionnaires fully completed | Completion of outcome measures | Baseline, 6 weeks, and 12 weeks post randomisation | Explore the acceptability of the outcome measures for clinicians and patients |
| **Intervention acceptability** |  |  | After intervention has concluded | Exploration of cancer journey, service ecology, role of early insomnia intervention and experiences of SRT or SHE |

## METHODS AND ANALYSIS

### Trial Design

This pilot, two-centre, two-arm, parallel group, block randomised (1:1) controlled trial will compare SRT with a SHE control condition. Both groups will continue to receive standard breast cancer treatment as usual (TAU). Consistent with the requirements of a pragmatic trial, there will be no limitations upon usual care for either group. In this way, the trial represents a comparison of SRT+SHE (+TAU) vs. SHE (+TAU), permitting clear judgements to be made regarding the relative clinical utility of SRT. Due to the nature of the SRT intervention, it is not possible to blind the therapists and patients to treatment allocation. However, the statistician will be blinded to treatment allocation (i.e., SRT or SHE) until after data analysis is complete. The trial is prospectively registered with ClinicalTrials.gov (identifier NCT06294041). Assessments will take place at baseline, 6 weeks, and 12 weeks post-randomisation (see Figure 1 for trial flowchart).

**Figure 1.**
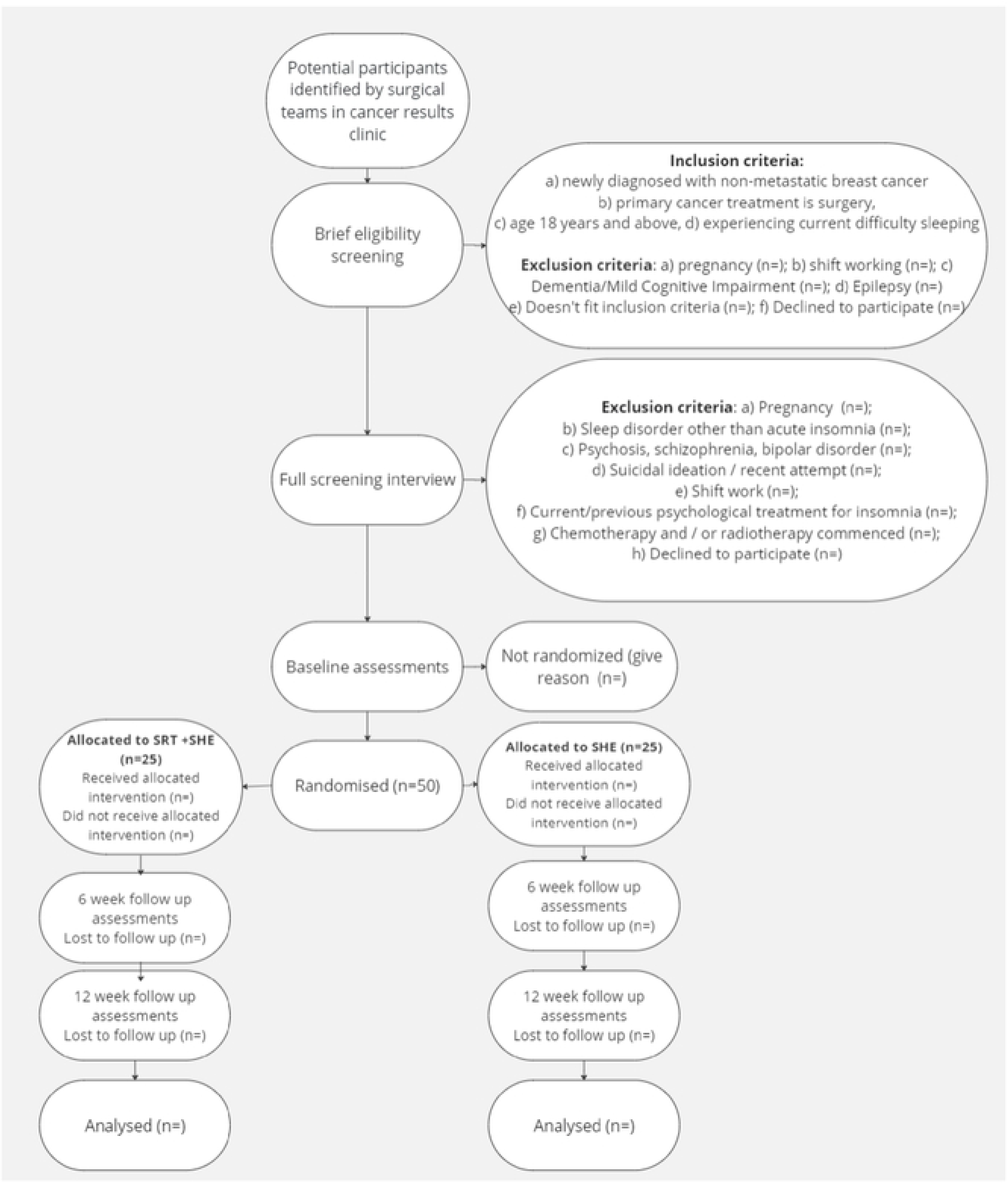
Trial flowchart UPLOADED SEPARATELY

### Recruitment

We will recruit 50 newly diagnosed, non-metastatic, breast cancer patients whose primary cancer treatment is surgery. Patients are eligible for recruitment at any point between cancer diagnosis and the onset of chemotherapy or radiotherapy, but efforts will be made to recruit prior to surgery where possible. Potentially eligible patients will be identified by surgical teams at NHS Greater Glasgow and Clyde and NHS Grampian breast cancer results clinics. If the patient expresses interest in participating, a member of their clinical team (i.e., nurse therapist) will conduct a brief eligibility screening questionnaire to confirm initial eligibility. See Figure 1 for full details of the exclusion criteria at this stage. Upon completion of the brief eligibility screener, contact details for potentially eligible patients will be passed to the researcher, who will share the patient information sheet, undertake consent procedures (written consent will be obtained via Qualtrics), and conduct a full screening interview to confirm eligibility.

#### Inclusion criteria

1. Patient is willing and able to give informed consent
2. Aged 18 years and above
3. Screen positive for acute insomnia, defined as dissatisfaction with sleep quality or duration, accompanied by other night / daytime symptoms, present for between 2 weeks and 3 months
4. Newly diagnosed with non-metastatic breast cancer
5. Primary cancer treatment is surgery

#### Exclusion criteria

1. Pregnancy
2. Additional sleep disorder diagnosis (e.g., restless legs syndrome, obstructive sleep apnoea, narcolepsy) or screen “positive” for additional sleep disorder at screening interview
3. Dementia / Mild Cognitive Impairment
4. Epilepsy
5. Psychosis (schizophrenia, bipolar disorder)
6. Current suicidal ideation with intent or attempted suicide within past 2 months
7. Night, evening, early morning, or rotating shiftwork
8. Current / previous psychological treatment for insomnia during the last 12 months
9. Chemotherapy and / or radiotherapy commenced

Screening will include an assessment of current sleep status to confirm the diagnosis of acute insomnia, using the Sleep Condition Indicator (18). It will also include a review of other relevant medical information (diagnosis of other sleep disorders, other medical and psychiatric disorders) to establish that there are no medical or psychiatric conditions that might preclude participation in the trial. Appropriate onward signposting will take place if any information is disclosed during screening that merits further investigation or follow-up (e.g. active suicidality). This procedure is made clear in the Patient Information Sheet.

### Interventions

Sleep Hygiene Education (SHE): SHE has been used as a control condition in many trials evaluating SRT. It does not have any therapeutic benefit for insomnia but is often offered as part of usual care, so is a credible alternative to SRT. SHE will be delivered via a booklet that provides information about lifestyle changes (e.g., having a light snack before bedtime, reducing caffeine) and changes to the bedroom environment (e.g., ensuring a dark room, comfortable mattress, optimal room temperature). Patients in the SHE condition will be asked to implement the SHE advice over a 4-week period. One week after randomisation to SHE, the researcher will telephone patients to check they understand the SHE advice and answer any questions they may have. The SRT nurse therapist will have no input into delivery of SHE and will not review patients randomised to this condition.

Sleep Restriction Therapy (SRT): SRT is a manualised, adaptive, behavioural insomnia intervention that is a key active ingredient within multi-component CBT-I. Our SRT protocol involves regularising and (where required) limiting a patient’s time in bed with the aim of increasing homeostatic sleep pressure, over-riding cognitive and physiological arousal, and strengthening circadian control of sleep (11). Those randomised to SRT will receive 4 weekly sessions totaling approximately 75 minutes of patient-therapist contact time (a detailed description of intervention content is provided in the TIDieR checklist in supplementary table 2). SRT will be administered by nurse therapists, trained by LF during a half-day training workshop. LF will also provide supervision to the nurse therapists throughout the intervention implementation phase as required. The nurse therapists will work collaboratively with the patients over a 4-week intervention phase to develop a tailored sleep/wake schedule, by restricting and adjusting time in bed where required. Patients randomised to SRT will also receive a SHE booklet (as above).

The four-week SRT intervention will be structured as follows:

- Session 1 (30 minutes) - The first session will be delivered face to face (either in person or online). The nurse therapist will introduce the rationale for SRT to the patient, review baseline sleep diary data, focus on core SRT principles (not extending time in bed, avoiding napping, keeping regular bed/rise times, maximising daytime natural light exposure, minimising nighttime artificial light exposure), and prescribe new bed and rise times if required. Advice on the management of daytime sleepiness and discussion of barriers/facilitators to implementation will also be offered. Patients will be given a workbook supporting home implementation of SRT and SHE guidance.
- Session 2 (15 minutes) - The nurse therapist will telephone the patient to check progress, provide an opportunity for questions, and advise on titration of the patient’s sleep schedule for the following week based on a structured algorithm.
- Session 3 (20 minutes) - The third session will follow the same structure as session two but will be a face-to-face session. The nurse therapist will review patient progress, explore (and attempt to overcome) barriers to implementation, and further titrate the sleep schedule (where required).
- Session 4 (10 minutes) - The nurse therapist will telephone the patient to check progress, provide an opportunity for questions, and advise on further titration of the patient’s sleep schedule if required. Suggestions for ongoing implementation and the management of residual or recurrent insomnia symptoms will also be provided during this final session.

### Assessments

Following completion of screening and consent procedures, eligible patients will be sent an email inviting them to complete a baseline (pre-intervention) assessment. Questionnaires can be completed online, over the telephone, or on paper, depending on patient preference. Insomnia severity will be assessed by the Insomnia Severity Index (ISI) (19), which is valid and sensitive to change in insomnia severity in cancer patients. We will also assess depressive symptoms [Patient Health Questionnaire (PHQ-9)] (20), anxiety symptoms [Generalised Anxiety Disorder Assessment (GAD-7)] (21), fatigue [Functional Assessment of Cancer Therapy (FACT-F)] (22), rest-activity rhythms [Actigraphy, CamNtech Ltd. MotionWatch 8], estimates of subjective sleep [Consensus Sleep Diary] (23), and health-related quality of life [EQ-5D-5L] (24) (from which, quality adjusted life years [QALYs] can be calculated). Within-treatment session outcomes for SRT will also be explored, so patients will be required to complete sleep diaries throughout the 4-week intervention phase providing information on sleep-wake cycles, such as bedtime, sleep onset latency, wake time after sleep onset, morning rise time, total sleep time, ratings of sleep quality and napping behaviour. After completion of baseline assessments, patients will be randomised and either referred to the nurse therapist for commencement of the SRT protocol or provided with the SHE resource. Following completion of the final assessment (12 weeks post-randomisation), those randomised to the SHE condition will be offered the opportunity to complete the SRT protocol without the requirement to complete any further assessments.

### Randomisation and Blinding

Following completion of baseline assessments, patients will be block randomised (1:1) to either SRT or SHE. The block size will vary from 2 to 4. The randomisation procedure will be administered by the statistician (DY). Due to the nature of the SRT intervention, it is not possible to blind the researcher, nurse therapist, or patients to treatment allocation. However, the statistician will be blinded to treatment allocation until data analyses are complete.

### Sample Size

As our focus is on feasibility, this trial is not formally powered to test clinical effects. However, based on a mean difference in ISI scores of 4.2 between groups (11) and an estimated standard deviation of 4.5 (17), a sample size of 40 patients (20 in each arm) would be required to achieve a power of 80% with a 5% significance level. We plan to randomise 50 patients (25 to each trial arm) based on an assumed dropout rate of 20% post randomisation. The results from this feasibility trial will help to refine the sample size calculation for a future definitive RCT.

### Qualitative Process Evaluation

To explore both trial and intervention processes, we will qualitatively explore contextual factors, implementation procedures, and intervention mechanisms of action to fully understand how nurse administered SRT is provided and received in oncology settings. This work complements the quantitative success criteria outlined in Table 2. Specifically, we will evaluate how intervention training is delivered to nurses and its acceptability to them, how the intervention is administered by nurses and whether intervention fidelity is maintained, the patient’s response to the intervention, how they interact with it, what they do as a result and any unexpected consequences. We will also explore any contextual factors within clinics and the wider health service environment that affect intervention implementation during the trial, increasing understanding of barriers and facilitators to intervention implementation, and providing insights for a future trial. To explore these issues, in-depth semi-structured interviews / focus groups (in-person, online, telephone) will be conducted by the researcher across both sites, and we will use theoretical frameworks (25, 26) designed explicitly to understand implementation processes. The sample size for this qualitative process evaluation is 20, comprising 15 patients (10 from SRT, 5 from SHE) and 5 clinical staff (including the nurse therapist and those responsible for patient recruitment).

### Adverse Events and Criteria for Discontinuation

Adverse Events (AEs) are defined as any unwanted medical event, reaction or side effect that occurs during the trial. AEs unrelated and related to the interventions will be recorded during treatment. Relatedness will be assessed by the researcher in the first instance, who will complete an adverse event reporting form. This form will then be reviewed and signed off by the CI. An AE will be classified as an unanticipated problem (UP), if the AE is unexpected, if it is related or possibly related to the trial procedures, and if the occurrence indicates that the patients or others are placed at greater risk by completing the trial than previously assumed. Serious Adverse Events (SAEs) such as deaths, suicide attempts, and serious accidents will also be recorded, captured via patient report and/or clinical records. We will also encourage patients to notify the research team of any change to their health and wellbeing that occur during the trial, in order that these can be recorded.

All AEs will be recorded in an adverse event reporting form, where a detailed description of the event and follow-up action will be noted. All SAEs that are deemed to be related to the trial will be reported to the Ethics Committee within 48hrs, and UPs that do not meet criteria for SAEs will be reported to the Chief Investigator within 48hrs, who is responsible to reporting to the Ethics Committee within 30 days. All remaining AEs will be collated for review by the trial steering group (comprised of four independent members, the trial researcher, and the CI).

This trial will be stopped prior to completion if: (a) the intervention is associated with AEs that call into question the safety of the intervention (> 20% of patients develop a SAE related to the intervention); or (b) any new information becomes available during the trial that necessitates stopping the trial; or (c) other situations occur that might warrant stopping the trial. Given previous research on SRT in breast cancer, it is unlikely that these thresholds will be met in this trial. Any decisions regarding discontinuation of the intervention would be made in consultation with the REC and the trial steering group.

### Data Analysis

As this is a pilot trial, interpretation of the results will be considered with reference to the MRC Framework for Complex Interventions. Details of patient screening, recruitment, retention, withdrawal, adherence, and follow-up will be reported descriptively as outlined in Table 2 above. To assess intervention acceptability, focus group/interview data will be transcribed and analysed using a Framework approach for qualitative data analysis supported by QSR NVivo. Analysis will occur as the interviews are transcribed and we will analyse qualitative process data prior to knowing trial outcomes to avoid biased interpretation. To fulfil our secondary aim, between group comparisons of ISI 6-weeks post-randomisation will be done using t-tests or Mann-Whitney tests. Effect sizes, along with 95% confidence intervals, will be reported for per-protocol and intention to treat analysis.

### Public and Patient Involvement (PPI)

In preparation for commencement of this trial, we recruited 5 members of our (previously established) PPI network to help shape the research questions, develop the project design, select suitable outcome measures, advise on the recruitment and data collection strategy, and contribute to proposal writing and editing. This ensures that patient wellbeing and improved patient care are central to our methodology. PPI colleagues will be invited to join each Investigator meeting (every 3 months during the trial), and one is a co-author on this paper. Our PPI network will also help to inform our wider dissemination and communication strategy by identifying key groups who will benefit most from trial outcomes.

### Data Management

Data will be managed in accordance with GDPR. The data management plan (DMP) is stored at the University of Strathclyde’s DMP repository according to University’s Research Data Management Policy and is subject to regular reviews. Data monitoring will be handled by the independent trial steering committee. After data collection is complete, de-identified data from this trial will be made available as open data through a research data repository [e.g., National Sleep Research Resource, https://sleepdata.org/) and metadata on Pure. None of the recordings from the focus groups/interviews will be shared as open data. Request for data use will be approved by the CI. We will not approve use for commercial purposes.

### Trial Status

The trial will open for recruitment in June 2024 and will continue recruiting until approximately March 2025, with outcome data expected around April 2026. This trial has received favourable opinion from West of Scotland REC 3 (reference 23/WS/0113).

## Data Availability

No datasets were generated or analysed during the current study. All relevant data from this study will be made available upon study completion.

## Authors’ Contributions

LF is the Chief Investigator, conceived the project, had overall responsibility for the trial design and treatment design and drafted the trial protocol. BE, JM, SDK, KB, DY, VG contributed to trial design. SDK contributed to treatment design. BE/JM are responsible for patient identification. DY is responsible for statistical analysis. KB is responsible for economic analysis. LF/SZ are responsible for the process evaluation analysis. All authors inputted to the trial protocol and commented on the manuscript.

---

**Supplementary Table 1. SPIRIT checklist**

**Supplementary Table 2: Template for Intervention Description and Replication (TIDieR) checklist**

## References

1. Sivertsen B, Hysing M, Harvey AG, Petrie KJ. The Epidemiology of Insomnia and Sleep Duration Across Mental and Physical Health: The SHoT Study. Frontiers in Psychology. 2021;12.

2. Parthasarathy S, Vasquez MM, Halonen M, Bootzin R, Quan SF, Martinez FD, et al. Persistent insomnia is associated with mortality risk. Am J Med. 2015;128(3):268–75.e2.

3. Harrold EC, Idris AF, Keegan NM, Corrigan L, Teo MY, O’Donnell M, et al. Prevalence of Insomnia in an Oncology Patient Population: An Irish Tertiary Referral Center Experience. Journal of the National Comprehensive Cancer Network. 2020;18(12):1623–30.

4. Sharma N, Hansen CH, O’Connor M, Thekkumpurath P, Walker J, Kleiboer A, et al. Sleep problems in cancer patients: prevalence and association with distress and pain. Psychooncology. 2012;21(9):1003–9.

5. Riemann D, Krone LB, Wulff K, Nissen C. Sleep, insomnia, and depression. Neuropsychopharmacology. 2020;45(1):74–89.

6. Spielman AJ, Caruso LS, Glovinsky PB. A behavioral perspective on insomnia treatment. Psychiatr Clin North Am. 1987;10(4):541–53.

7. Fleming L, Randell K, Stewart E, Espie CA, Morrison DS, Lawless C, et al. Insomnia in breast cancer: a prospective observational study. Sleep. 2019;42(3).

8. Reynolds-Cowie P, Fleming L. Living with persistent insomnia after cancer: A qualitative analysis of impact and management. British journal of health psychology. 2021;26(1):33–49.

9. Fleming L, Gillespie S, Espie CA. The development and impact of insomnia on cancer survivors: a qualitative analysis. Psycho-Oncology. 2010;19(9):991–6.

10. Rehman A, Drake CL, Shiramizu V, Fleming L. Sleep reactivity predicts insomnia in patients diagnosed with breast cancer. Journal of Clinical Sleep Medicine. 2022:jcsm. 10170.

11. Maurer LF, Schneider J, Miller CB, Espie CA, Kyle SD. The clinical effects of sleep restriction therapy for insomnia: A meta-analysis of randomised controlled trials. Sleep Med Rev. 2021;58:101493.

12. Fleming L, Randell K, Harvey CJ, Espie CA. Does cognitive behaviour therapy for insomnia reduce clinical levels of fatigue, anxiety and depression in cancer patients? Psycho-Oncology. 2014;23(6):679–84.

13. Espie CA, Fleming L, Cassidy J, Samuel L, Taylor LM, White CA, et al. Randomized controlled clinical effectiveness trial of cognitive behavior therapy compared with treatment as usual for persistent insomnia in patients with cancer. Journal of Clinical Oncology. 2008;26(28):4651–8.

14. Maurer LF, Ftouni S, Espie CA, Bisdounis L, Kyle SD. The acute effects of sleep restriction therapy for insomnia on circadian timing and vigilance. J Sleep Res. 2021;30(4):e13260.

15. Maurer LF, Espie CA, Omlin X, Reid MJ, Sharman R, Gavriloff D, et al. Isolating the role of time in bed restriction in the treatment of insomnia: a randomized, controlled, dismantling trial comparing sleep restriction therapy with time in bed regularization. Sleep. 2020;43(11).

16. Palesh O, Scheiber C, Kesler S, Janelsins MC, Guido JJ, Heckler C, et al. Feasibility and acceptability of brief behavioral therapy for cancer-related insomnia: effects on insomnia and circadian rhythm during chemotherapy: a phase II randomised multicentre controlled trial. British journal of cancer. 2018;119(3):274–81.

17. Kyle SD, Siriwardena AN, Espie CA, Yang Y, Petrou S, Ogburn E, et al. Clinical and cost-effectiveness of nurse-delivered sleep restriction therapy for insomnia in primary care (HABIT): a pragmatic, superiority, open-label, randomised controlled trial. Lancet (London, England). 2023;402(10406):975–87.

18. Espie CA, Kyle SD, Hames P, Gardani M, Fleming L, Cape J. The Sleep Condition Indicator: a clinical screening tool to evaluate insomnia disorder. Bmj Open. 2014;4(3).

19. Bastien CH, Vallières A, Morin CM. Validation of the Insomnia Severity Index as an outcome measure for insomnia research. Sleep Med. 2001;2(4):297–307.

20. Kroenke K, Spitzer RL, Williams JB. The PHQ-9: validity of a brief depression severity measure. J Gen Intern Med. 2001;16(9):606–13.

21. Spitzer RL, Kroenke K, Williams JB, Löwe B. A brief measure for assessing generalized anxiety disorder: the GAD-7. Arch Intern Med. 2006;166(10):1092–7.

22. Yellen SB, Cella DF, Webster K, Blendowski C, Kaplan E. Measuring fatigue and other anemia-related symptoms with the Functional Assessment of Cancer Therapy (FACT) measurement system. J Pain Symptom Manage. 1997;13(2):63–74.

23. Carney CE, Buysse DJ, Ancoli-Israel S, Edinger JD, Krystal AD, Lichstein KL, et al. The consensus sleep diary: standardizing prospective sleep self-monitoring. Sleep. 2012;35(2):287–302.

24. Herdman M, Gudex C, Lloyd A, Janssen M, Kind P, Parkin D, et al. Development and preliminary testing of the new five-level version of EQ-5D (EQ-5D-5L). Quality of life research: an international journal of quality of life aspects of treatment, care and rehabilitation. 2011;20(10):1727–36.

25. Nilsen P. Making sense of implementation theories, models and frameworks. Implementation Science. 2015;10(1).

26. Moore GF, Audrey S, Barker M, Bond L, Bonell C, Hardeman W, et al. Process evaluation of complex interventions: Medical Research Council guidance. Bmj. 2015;350:h1258.

